## Supplementary figures and images for "Machine learning detects hidden treatment response patterns only in the presence of comprehensive clinical phenotyping"

### Supplementary information

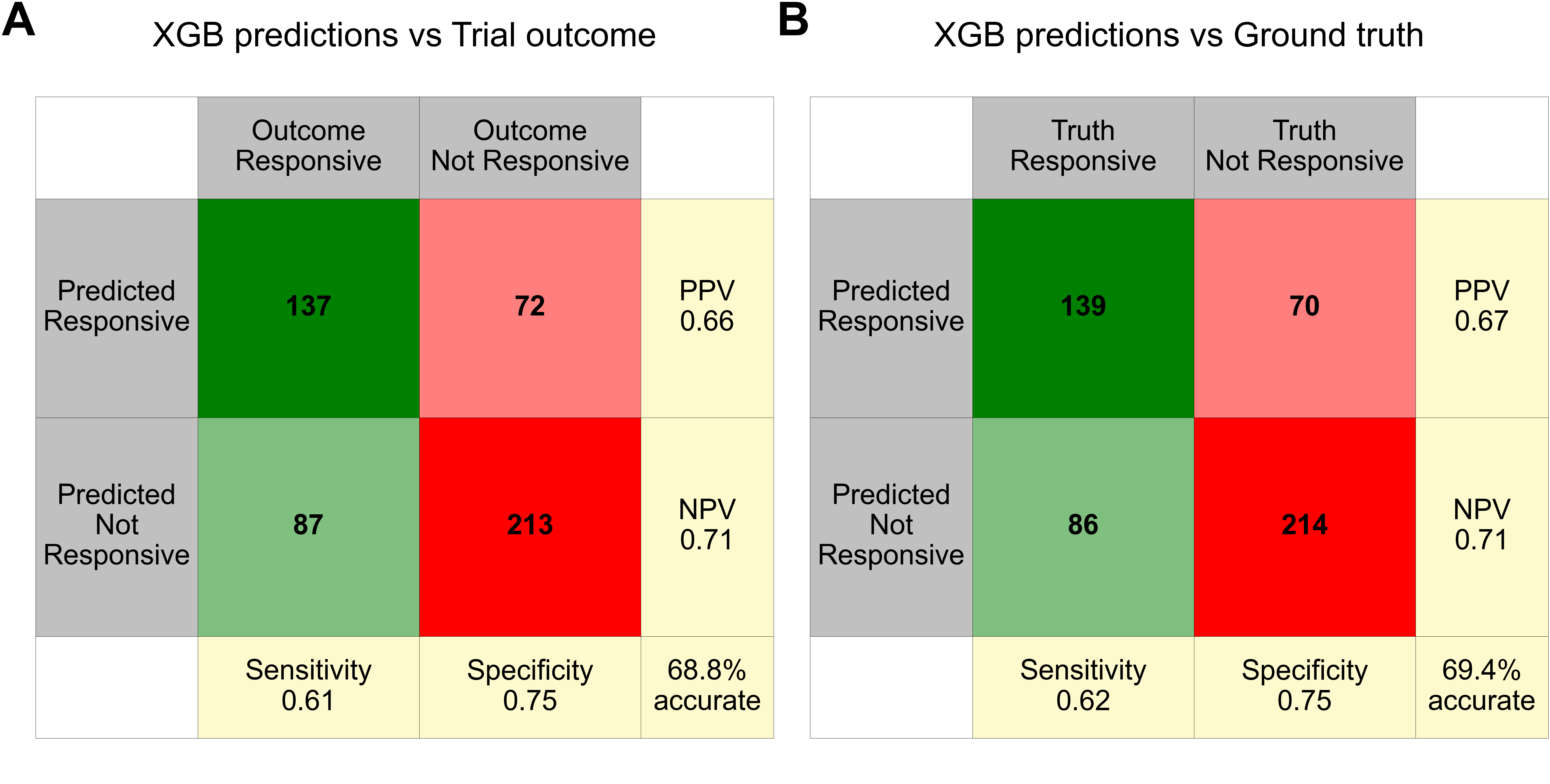

**Supplementary Figure 1**
